# Understanding men’s participation in mass drug administration: evidence from a cluster-randomised trial of community-based deworming in Malawi

**DOI:** 10.1101/2025.11.21.25340746

**Authors:** Stefan Witek-McManus, James Simwanza, Rejoice Msiska, Alvin Chisambi, Sean R. Galagan, William E. Oswald, Elliott Rogers, Joseph Timothy, Hugo Legge, Judd L. Walson, Lazarus Juziwelo, Rachel L. Pullan, Robin L. Bailey, Khumbo Kalua

**Affiliations:** Department of Disease Control, Faculty of Infectious and Tropical Diseases, London School of Hygiene & Tropical Medicine, London, UK; Blantyre Institute for Community Outreach, Blantyre, Malawi; The DeWorm3 Project, University of Washington, Seattle, USA; Department of International Health, Bloomburg School of Public Health, John Hopkins University, Baltimore, USA; STH and Schistosomiasis Control Programme, Community Health Sciences Unit, Ministry of Health & Population, Lilongwe, Malawi; Department of Clinical Research, Faculty of Infectious and Tropical Diseases, London School of Hygiene & Tropical Medicine, London, UK; Kamuzu University of Health Sciences, Blantyre, Malawi; School of Population and Public Health, University of British Columbia, Vancouver, Canada

**Keywords:** Malawi, mass drug administration, men’s health, neglected tropical diseases, soil-transmitted helminths

## Abstract

1.

**Background:** Mass drug administration (MDA) is a key strategy in the response to neglected tropical diseases (NTDs). Operational research indicates that treatment of adult men during MDA is frequently lower than that of adult women, but this remains poorly understood and obscured by a lack of routine sex-disaggregated reporting. Such differences may represent a potential threat to NTD strategies that require high MDA treatment coverage to achieve control or elimination objectives.

**Methods:** Using detailed treatment data from a cluster-randomised trial of community-based MDA for soil-transmitted helminths (STH) conducted in southern Malawi, this secondary analysis describes sex-specific trends in coverage over six consecutive rounds of MDA delivered biannually at household level. We report community-level coverage by both a per-protocol (trial) definition, and a pragmatic operational definition to simulate routine implementation. We used multilevel mixed-effects logistic regression to investigate individual, household and programme factors associated with non-treatment by both coverage definitions amongst adult men.

**Results:** Adult men’s participation in MDA was substantially and consistently lower than adult women. Median difference (women minus men) in community-level protocol coverage was 15.7% (IQR 10.4–21.3%), increasing to 23.2% (IQR 17.5–30.2%) by operational coverage. Factors associated with increased odds of non-treatment were similar by coverage definition and included younger age, lower levels of education, and previous reported absenteeism; and were reduced by modifiable programmatic factors such as the day or time of household visit.

**Conclusions:** This analysis highlights the persistence of inequity for adult men in participation in MDA, despite a well-resourced context that achieved high coverage of adult women. More nuanced MDA strategies that respond to the needs of adult men will be required to achieve equitable coverage, accompanied by robust monitoring and evaluation to evaluate their effectiveness at reaching this under-served population.

**What is already known on this topic:**

- Achieving high coverage of mass drug administration (MDA) across population groups is key to targets for control and elimination of many neglected tropical diseases (NTDs) and achieving global health equity.
- Men’s lower participation in MDA has been recognised by routine programmes but has received limited research and policy attention, compounded by a lack of sex-disaggregated monitoring and evaluation.

**What this study adds:**

- We show that high overall levels of MDA coverage masked substantial inequity, with adult men’s coverage in MDA substantially and consistently lower than women.
- We identify specific factors associated with non-treatment of adult men – including modifiable programmatic characteristics – that could be targeted by programmes seeking to address this disparity.

**How this study might affect research, practice or policy:**

- These findings underscore the need for MDA strategies to comprehensively assess and address coverage gaps among adult men, and should encourage policymakers to ensure that the design and implementation of community-based NTD strategies reflect the needs of both men and women.

## 2. INTRODUCTION

Neglected tropical diseases (NTDs) are a diverse group of twenty diseases that disproportionately affect low and middle-income countries (LMICs). Estimated to have caused 10.3 to 25.3 million disability adjusted life years (DALYs) and 65 to 143 thousand deaths globally in 2021 [1], almost 1.5 billion people required mass or individual treatment for NTDs in 2024 [2]. A core strategic intervention for the control or elimination for five NTDs – lymphatic filariasis, onchocerciasis, schistosomiasis, soil-transmitted helminths (STH) and trachoma – is the use of mass drug administration (MDA), whereby a therapeutic dose of medicine is routinely delivered to target populations at scale [3]. Key to the success of MDA is ensuring that the specific delivery approach achieves treatment of a high proportion, or coverage, of the target population, while also maximizing programme efficiency and equity across socio-demographic factors [4].

Current global targets specifically prioritise control of soil-transmitted helminths (STH), through reduction of morbidity amongst specific populations at greatest risk of anaemia and impaired nutrition caused by STH infection, including pre-school children (PSAC), school-aged children (SAC), and women of reproductive age (WRA) [5]. With a target of 75% coverage amongst those at risk, MDA for STH is generally delivered through platforms that can efficiently reach these populations at scale: by teachers through schools; or in combination with pre-existing services, such as by community health workers during community outreach and antenatal visits [6, 7]. Supported by large donations of drugs and sustained external investment, MDA for STH has come to be one of the largest public health interventions globally, reaching 492 million people in need in 2022 [8].

Having consistently achieved high levels of coverage of MDA for STH amongst at-risk groups and concomitant reductions in STH prevalence, increasing attention has been placed on the feasibility of eliminating STH infection entirely [9]. Such strategies would require expanding those targeted by MDA to all eligible members of a community, who are likely to be significant reservoirs of infection and therefore key to interrupting STH transmission; and in consistently achieving high levels of MDA coverage amongst the broader community [10]. This expanded target population would most significantly include the addition of adult men, a population for which low coverage of MDA for other NTDs has previously been recognized, but which has remained largely unaddressed in programme delivery [11–13].

Better understanding of the barriers that men face in accessing health services, and addressing areas where men’s health outcomes are worse, remains conspicuously absent from the global health agenda [14]. Given that PSAC, SAC and WRA remain the priority groups for STH control, it is not surprising that in a variety of settings, prevalence and intensity of STH infection remains highest amongst adult men [15–17], but evaluation of the feasibility of STH elimination presents greater urgency to understanding men’s participation in MDA: poor coverage may affect not only the (worse) health of men, but present a broader risk to the health of women and children through a persistent reservoir of infection in their community, compromising elimination efforts.

Opportunities to examine socio-demographic differences in MDA coverage amongst adults are surprisingly rare, in part due to a lack of disaggregated reporting and evaluation [18, 19]. This study uses individual-level treatment data from the DeWorm3 trial, a multi-site cluster-randomised trial in Benin, Malawi and India that evaluated the feasibility of eliminating STH through biannual community-wide deworming (cMDA) [20]. A baseline survey of the Malawi site in 2018 confirmed that STH prevalence was highest amongst adult men [21]. A cross-site analysis of cMDA reported that while study cluster-level aggregated coverage was generally high over six consecutive rounds, women were significantly more likely to have been treated as compared to men, most notably in the Malawi trial site [22]. The objectives of this analysis were therefore to (i) explore differences in cMDA coverage between adult men and women over time, and to (ii) identify specific factors associated with non-treatment of adult men during cMDA in Malawi.

## 3. METHODS

### 3.1. Study design and participants

The DeWorm3 trial design (protocol) [20] and intervention coverage [22] across all three trial sites have previously been described, in addition to a baseline survey [21] and descriptions of both standard of care (control) and intervention arms [23] in Malawi. In brief, 124 communities (gazetted villages or towns) in Namwera Health Zone in Mangochi district were assigned to 40 study clusters, and clusters subsequently randomised 1:1 to two specific types of MDA using restricted randomisation: annual school-based deworming (SBD) (national standard of care) of children aged 5–14 years of age, or annual SBD plus biannual cMDA (intervention) of all censused individuals ≥2 years of age. The trial primary outcome was transmission interruption, defined as cluster-level population prevalence of any STH species ≤2% by quantitative PCR (qPCR), following six biannual rounds of cMDA over three years and a further two years of no cMDA. Written informed consent was obtained from an adult household member for participation in the annual community census, and verbally (per national guidelines) for participation in the intervention at each round of cMDA. The DeWorm3 trial was approved by the College of Medicine Research Ethics Committee at the University of Malawi (P.04/17/2161), the London School of Hygiene and Tropical Medicine (LSHTM) Observational/Interventions Research Ethics Committee (12013), and the Human Subjects Division at the University of Washington (STUDY00000180). The results of this study are reported in-line with Sex And Gender Equity in Research (SAGER) guidelines [24].

### 3.2. Procedures

An annual community-based census was conducted in all study clusters between November-December 2017 (baseline census), March-May 2019 (census update 1) and April-July 2020 (census update 2). Trained study enumerators visited all known households within a defined community with the support of volunteer village guides. All occupied and consenting households completed a census questionnaire administered by the enumerator, which comprised a demographic listing of all household members, reported household-level demographic characteristics, observations of dwelling construction materials, and reported access to water and sanitation facilities. Subsequent census questionnaires to a household comprised only of reviewing the household listing and updating household members (births, deaths and inward or outward migration).

In intervention clusters, cMDA was delivered biannually between 2018–2020 by a cadre of community health workers known as Health Surveillance Assistants (HSAs), who were responsible for delivery of cMDA to residents within their routine catchment areas. cMDA was primarily delivered ‘door-to-door’, where an HSA (supported by a study officer and community volunteer) offered treatment with a single dose of 400mg albendazole to all resident and eligible household members and to directly observe treatment, although HSAs were permitted to leave tablets with an individual if requested, or with another present household member if a resident was absent at a third consecutive visit (i.e. treatment not directly observed). Recording of treatment was completed by the accompanying study officer using an electronic treatment register, pre-populated with a household and individual listing based on the most recent census [25]. Real-time monitoring and supervision of cMDA was conducted by the trial study team, which included providing summary coverage statistics to HSAs and generating ‘mop-up’ lists of non-treated community members to be targeted during the cMDA delivery period.

### 3.3. Definitions and outcomes

We defined the population targeted for cMDA (denominator) as adults (≥18 years of age) registered or confirmed at the most recent census prior to cMDA and recorded as eligible (i.e. not pregnant in first trimester and not deceased) at that round of cMDA. Consistent with the primary coverage definition used in the parent trial, we first defined *protocol* treatment as those recorded as receiving treatment whether directly or not directly observed (tablet left with individual, tablet left with other household member, treated elsewhere within past two weeks). Considering the trial context in which cMDA was delivered, we additionally defined *operational* treatment using a pragmatic definition that better reflects how cMDA would likely be delivered in routine practice, and which aligns more closely with how cMDA for other NTDs is delivered in Malawi: those recorded as receiving any treatment (observed or unobserved) at the *first* visit to their household (therefore with those treated at a second or third visit, or for whom tablets were left with another household member, considered not treated). Irregular residents were defined as individuals who were reported at the census to be household members but had not been present for the majority of days in the prior six months. Household urbanicity (rural versus peri-urban) was *a priori* defined based on relative characteristics of each study community and incorporated both local facilities (e.g. presence of a trading centre) and spatial (e.g. household density) characteristics.

### 3.4. Statistical analysis

Analysis was conducted using Stata 18.5 (StataCorp, TX, USA). Household socio-economic status (SES) was calculated using principal component analysis (PCA) as previously described [21]. Community-level coverage of cMDA is described using descriptive statistics (median (M), interquartile range (IQR)) and presented by sex for cMDA overall and by round. Differences in community-level coverage of cMDA were calculated as the median difference and IQR between communities by round. We apply a target of 75% cMDA coverage for STH, based on similar current global targets for routine MDA for STH. Consistent with a multi-level (clustered) dataset, we conducted mixed-effects logistic regression fitted by maximum likelihood estimation to investigate individual, household and programme factors associated with non-treatment amongst adult men across all six rounds of cMDA. Iterative versions of models were tested with variance partition coefficient calculated to assess statistical significance of the clustering by a likelihood ratio test, a summary of this is reported in Supplementary Table 3. The optimal model structure was identified as a three-level hierarchy with treatment at each round (level-1) nested within individual (level-2), in turn nested within community (level-3). The final model comprised a random intercept for individual and community to allow for baseline variation in non-treatment at each of these levels; and a random slope for cMDA round at the individual level, to allow for the reduction in non-treatment amongst individuals across rounds (i.e. over time). The effect of contextual variables treated as fixed effects included those from the relevant (i.e. previous to that round of cMDA) census record (age, education, absenteeism, SES, household size, religion, language, urbanicity) or cMDA treatment record (time and day of treatment) and were all pre-specified to be included in an adjusted model.

## 4. RESULTS

### 4.1. Summary cMDA treatment outcomes, by sex and cMDA round

A summary of cMDA outcomes by sex and round is presented in Table 1. The proportion of adult men with no household visit recorded (i.e. treatment not offered) ranged between 2.4% (round 5) and 10.4% (round 2), and was higher than amongst adult women at every round. A greater proportion of adult men were recorded as absent with substantial variation by round, ranging from 11.5–17.2% amongst those reported to be short term absent, and 2.5–7.8% for those reported to be long term absent; in contrast, the proportion of adult women reported as short-term absent (2.4–3.7%) and long-term absent (0.8–2.0%) remained consistent and low across rounds. Amongst adults reached but not treated, the most common reason amongst men was refusal (range by round: 0.1–3.2%) whereas amongst women it was non-eligibility (range by round: 1.5–2.5%). The proportion of treatments administered without direct observation generally declined over each round but was always higher amongst adult men (range by round: 1.4–11.5%) than adult women (range by round: 0.7–5.4%).

**Table 1:** Summary treatment outcomes amongst censused adults during cMDA, by sex and round.

|  | cMDA 1 |  | cMDA 2 |  | cMDA 3 |  |
| --- | --- | --- | --- | --- | --- | --- |
|  | Adult men<br>n= (%) | Adult women<br>n= (%) | Adult men<br>n= (%) | Adult women<br>n= (%) | Adult men<br>n= (%) | Adult women<br>n= (%) |
| **Censused (N=)** | **12 061** | **15 563** | **12 391** | **15 655** | **11 958** | **15 768** |
| **Not reached:** |  |  |  |  |  |  |
| Not visited | 1 064 (8.8) | 1 165 (7.5) | 1 288 (10.4) | 1 435 (9.2) | 406 (3.4) | 455 (2.9) |
| Short-term absent | 1 792 (14.9) | 466 (3.0) | 2 126 (17.2) | 579 (3.7) | 1 885 (15.8) | 479 (3.0) |
| Long-term absent | 936 (7.8) | 312 (2.0) | 652 (5.3) | 267 (1.7) | 395 (3.3) | 133 (0.8) |
| **Not treated:** |  |  |  |  |  |  |
| Not eligible<sup>†</sup> | 67 (0.6) | 275 (1.8) | 106 (0.9) | 392 (2.5) | 28 (0.2) | 312 (2.0) |
| Refused | 383 (3.2) | 132 (0.9) | 246 (2.0) | 85 (0.5) | 146 (1.2) | 26 (0.2) |
| Other<sup>‡</sup> | 11 (0.1) | 24 (0.2) | 41 (0.3) | 73 (0.5) | 53 (0.4) | 52 (0.3) |
| **Treated:** |  |  |  |  |  |  |
| Observed | 6 482 (53.7) | 12 521 (80.5) | 6 513 (52.6) | 11 981 (76.5) | 8 233 (68.9) | 13 813 (87.6) |
| Not observed<sup>*</sup> | 1 326 (11.0) | 668 (4.3) | 1 419 (11.5) | 843 (5.4) | 812 (6.8) | 498 (3.2) |
|  | cMDA 4 |  | cMDA 5 |  | cMDA 6 |  |
|  | Adult men<br>n= (%) | Adult women<br>n= (%) | Adult men<br>n= (%) | Adult women<br>n= (%) | Adult men<br>n= (%) | Adult women<br>n= (%) |
| **Censused (N=)** | **12 379** | **16 119** | **12 561** | **16 330** | **13 383** | **16 804** |
| **Not reached:** |  |  |  |  |  |  |
| Not visited | 512 (4.1) | 604 (3.7) | 300 (2.4) | 345 (2.1) | 463 (3.5) | 548 (3.3) |
| Short-term absent | 1 578 (12.7) | 493 (3.1) | 1 442 (11.5) | 380 (2.3) | 1 727 (12.9) | 554 (3.3) |
| Long-term absent | 626 (5.1) | 269 (1.7) | 316 (2.5) | 164 (1.0) | 597 (4.5) | 265 (1.6) |
| **Not treated:** |  |  |  |  |  |  |
| Not eligible<sup>†</sup> | 83 (0.7) | 335 (2.1) | 23 (0.2) | 244 (1.5) | 62 (0.5) | 305 (1.8) |
| Refused | 54 (0.4) | 18 (0.1) | 9 (0.1) | 0 (0) | 5 (<0.1) | 3 (>0.1) |
| Other<sup>‡</sup> | 46 (0.4) | 55 (0.3) | 35 (0.3) | 52 (0.3) | 39 (0.3) | 61 (0.4) |
| **Treated:** |  |  |  |  |  |  |
| Observed | 8 853 (71.5) | 13 966 (86.6) | 10 256 (81.6) | 15 028 (92.0) | 10 102 (75.5) | 14 792 (88.0) |
| Not observed<sup>*</sup> | 627 (5.1) | 379 (2.4) | 180 (1.4) | 117 (0.7) | 388 (2.9) | 276 (1.6) |

\*: Tablet left with individual, tablet left with other individual in household, or treated elsewhere in previous two weeks. †: Pregnant in first trimester or deceased. ‡: Seriously ill or intoxicated.

### 4.2. Treatment coverage of cMDA: per-protocol (trial)

Across all six rounds of cMDA, median community-level protocol coverage of adult men was 74.1% (IQR 66.8–80.9%) compared to 90.7% (IQR 86.3–94.0%) of adult women. Median community-level protocol coverage of adult men was consistently lower compared to adult women at every round and remained below 75% during rounds 1 to 3, whereas community-level protocol coverage of adult women exceeded 75% in all 6 rounds (Figure 1).

**Figure 1:**
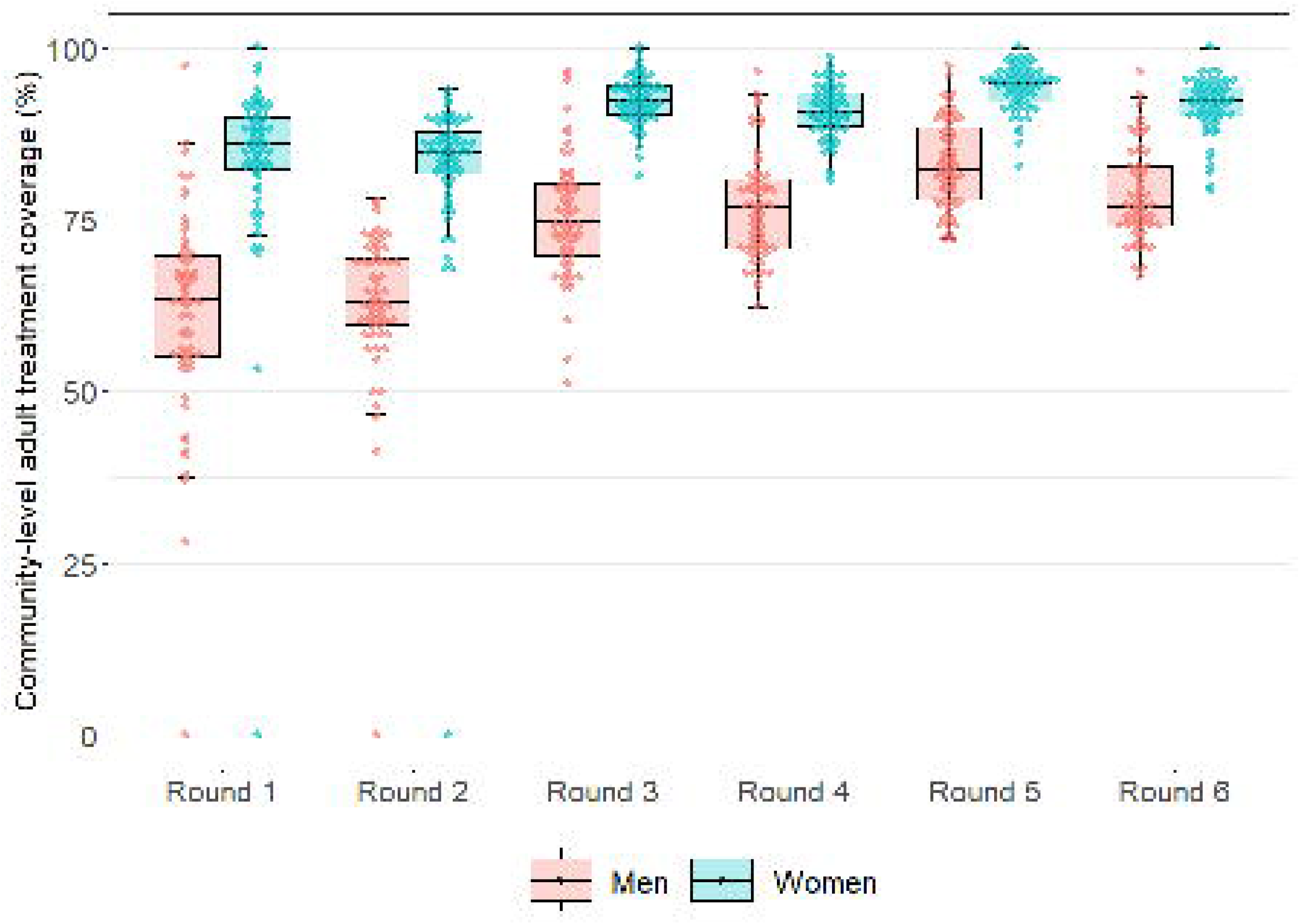
Community-level cMDA coverage of eligible adults, by sex and round: trial (per-protocol)

Median community-level protocol coverage of adult men was lowest at round 2 (62.9%, IQR 59.6–69.7%) and highest during round 5 (82.3%, IQR 77.8–89.1%), increasing each consecutive round from round 2 to 5. The overall proportion of communities with protocol coverage of adult men ≥75% was lowest during round two (3/60, 5.0%) and highest during round five (n=53/59, 89.8%). Median community-level protocol coverage of adult women was also lowest in round two (84.8%, IQR 81.3–88.2%) and highest in round five (94.9%, IQR 92.0–96.4%). The majority of communities achieved protocol coverage of adult women ≥75% during round 1 and round 2 (54/60, 90.0%), and in all communities during rounds 3 to 6.

### 4.3. Treatment coverage of cMDA: operational scenario

In comparison to protocol coverage, median community-level operational coverage of adult men overall across all six rounds of cMDA was substantially lower, and showed greater variation across communities (61.1%, IQR 51.3–68.9%) as compared to operational coverage of adult women (84.7%, IQR 79.7–89.3%). Median community-level operational coverage of adult men was below 75% in all six rounds, whereas operational coverage of adult women remained above 75% in all six rounds (Figure 2).

**Figure 2:**
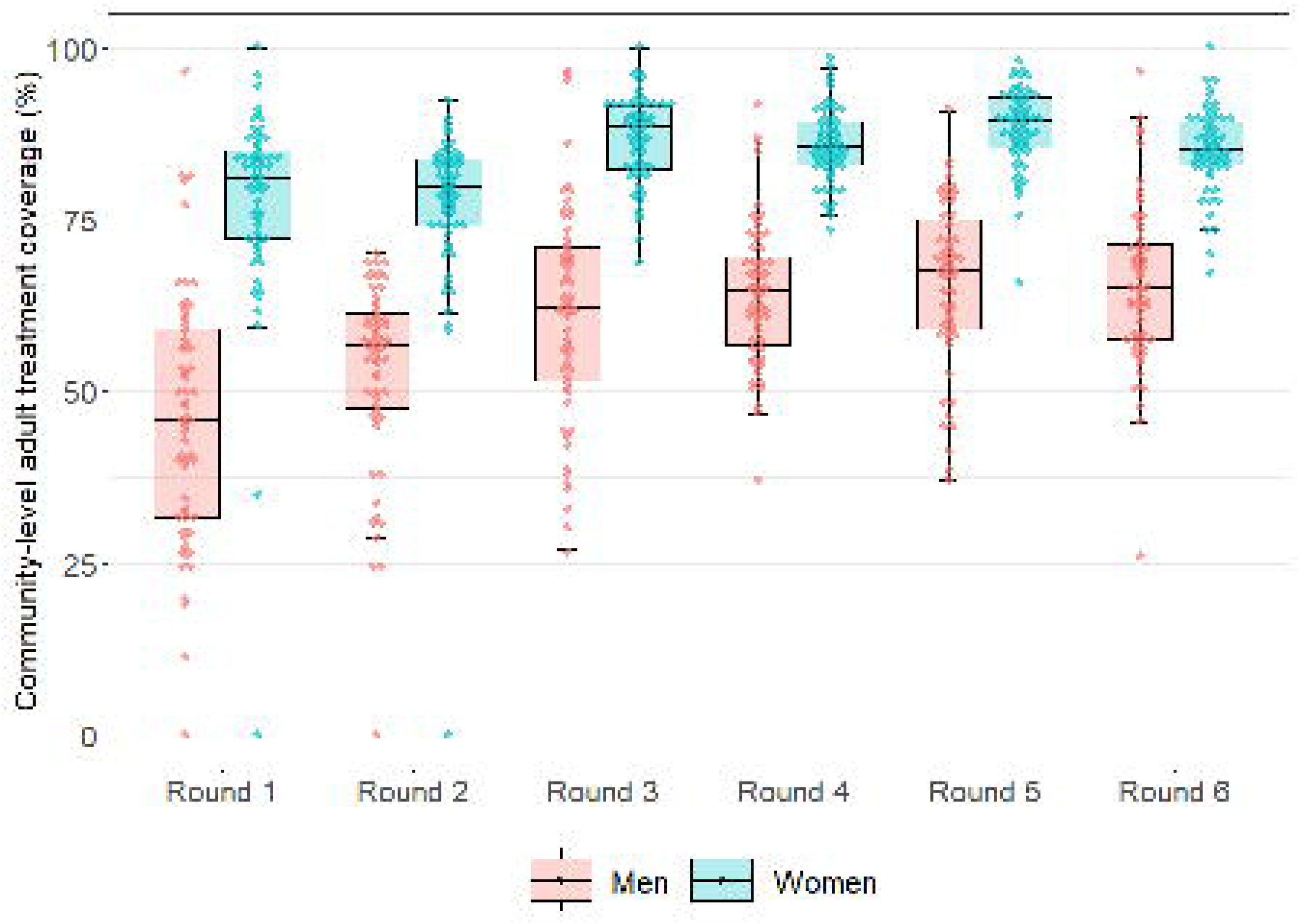
Community-level cMDA coverage of eligible adults, by sex and round: operational scenario

Median community-level operational coverage of adult men was lowest in round 1 (M=45.8%, IQR 31.6–59.2 %) and highest in round 5 (M=67.7%, IQR 58.8–75.5%). No community achieved per-protocol coverage of adult men ≥75% during round 2, with the highest proportion of communities achieving this (albeit substantially lower as compared to protocol coverage) during round 5 (15/60, 25.4%). In contrast to protocol coverage, in no round did all communities meet protocol coverage of adult women ≥75%, however, the majority did in round 4 and round 5 (n=58/59, 98.3%).

### 4.4. Within-community differences in cMDA treatment coverage

The median difference (adult women minus adult men) in community-level protocol coverage was 15.7% (IQR 10.4–21.3%) (Figure 3). Consistent with increasing coverage amongst adult men over successive cMDA rounds, median difference in protocol coverage was greatest in round 1 (M=22.2%, IQR 16.1–27.8%) and least in round 5 (10.8%, IQR: 6.2–15.6%). In every round, more than half of communities had a difference in coverage ≥10% (35/59–55/60, 59.3–91.7%) amongst adult women compared to adult men; and one or less communities where difference in protocol coverage was ever ≤0% (i.e. equal to or greater amongst adult men, as compared to adult women).

**Figure 3:**
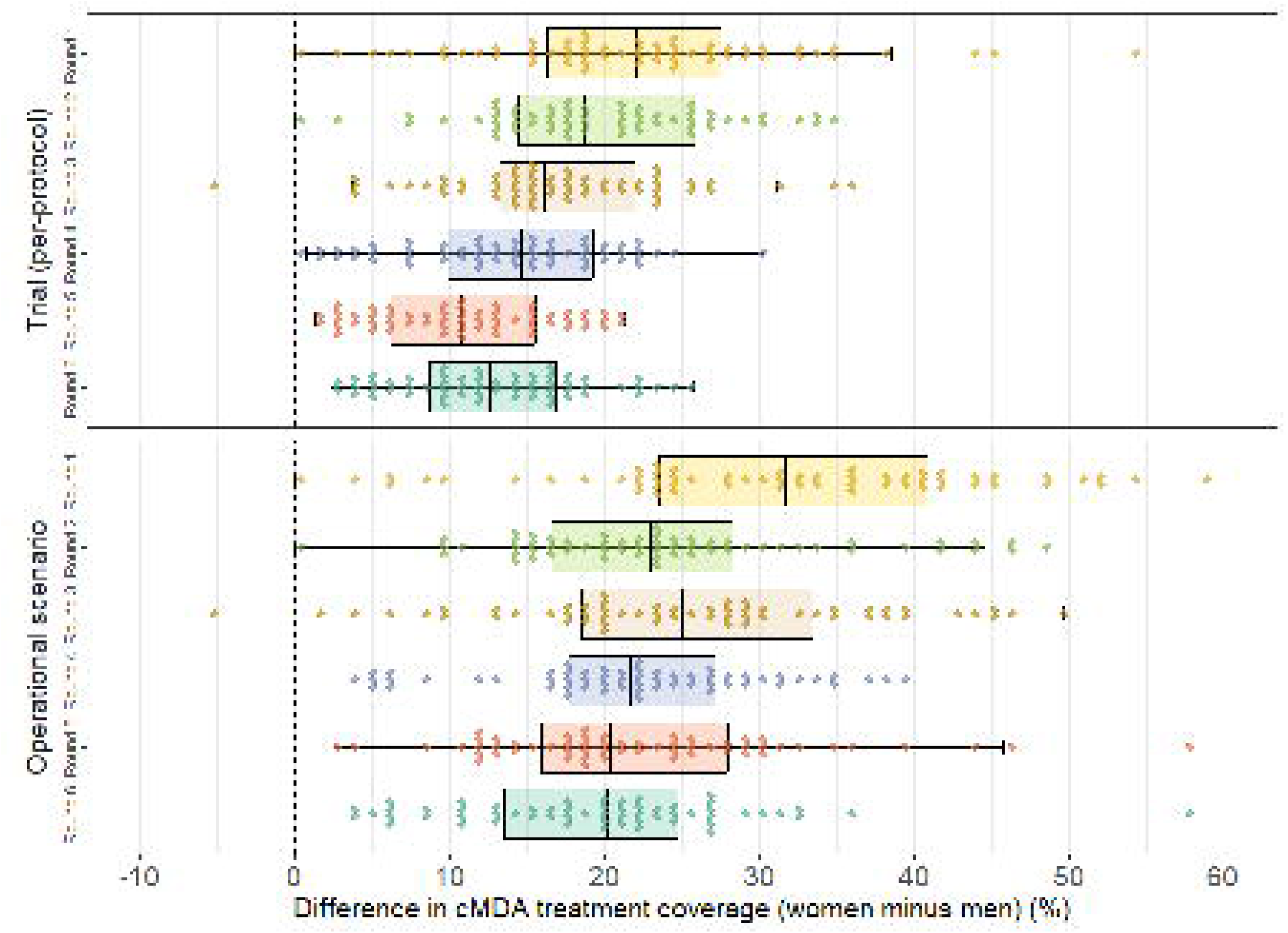
Within-community sex difference (women minus men) in community-level cMDA coverage of eligible adults, by coverage definition and cMDA round

The median difference in community-level operational coverage was 23.2% (IQR 17.5–30.2). In contrast to the difference in protocol coverage, change between rounds was inconsistent, and least in round 6 (M=20.3%, IQR 13.4–25.1%). However, the total proportion of communities with difference in coverage ≥10% amongst adult women compared to adult men remained high (320/356, 89.9%); with only ever one community where the difference in operational coverage was ≤0% (round 3 only).

### 4.5. Factors associated with non-treatment of adult men: per-protocol (trial) coverage

The results of multivariable modelling of individual, household and treatment-level characteristics of adult men and non-treatment across all 6 rounds of cMDA are presented for protocol non-treatment (Table 2) and for operational non-treatment (Table 3). Univariate associations are reported in Supplementary Table 1 (protocol (trial)) and Supplementary Table 2 (operational scenario).

**Table 2:** Proportion non-treated and factors associated with per-protocol (trial) non-treatment of censused and eligible adult men during cMDA.

| Factor | Number not treated<br>/total; n/N (%) | Adjusted OR<br>(95% CI)<sup>*</sup> | P-value |
| --- | --- | --- | --- |
| **Individual:** |  |  |  |
| **Age group:** |  |  |  |
| 18-19 years | 1 896/7 677 (24.7) | 1.0 | - |
| 20-24 years | 4 058/11 522 (35.2) | 1.58 (1.42, 1.74) | <0.001 |
| 25-29 years | 3 517/9 454 (37.2) | 1.71 (1.52, 1.91) | <0.001 |
| 30-39 years | 4 906/15 913 (30.8) | 1.20 (1.08, 1.33) | 0.001 |
| 40-49 years | 2 497/12 249 (20.4) | 0.65 (0.58, 0.73) | <0.001 |
| ≥50 years | 2 300/17 550 (13.1) | 0.38 (0.34, 0.42) | <0.001 |
| **Education level:** |  |  |  |
| None | 4 523/18 336 (24.7) | 1.0 | - |
| Primary incomplete | 7 494/25 408 (29.5) | 1.04 (0.97, 1.11) | 0.27 |
| Primary complete | 707/1 979 (35.7) | 1.19 (1.02, 1.38) | 0.03 |
| Secondary or higher | 2 211/6 710 (33.0) | 1.01 (0.91, 1.12) | 0.85 |
| Unknown | 4 239/21 932 (19.3) | 1.01 (0.92, 1.11) | 0.84 |
| **Presence in household** |  |  |  |
| Absent at census visit | 13 381/40 313 (33.2) | 1.23 (1.16, 1.30) | <0.001 |
| Absent night before census visit | 7 653/13 466 (56.8) | 3.00 (2.76, 3.25) | <0.001 |
| Irregular resident<sup>†</sup> | 5 697/ 9 046 (63.0) | 2.69 (2.45, 2.96) | <0.001 |
| **Household:** |  |  |  |
| **Wealth quintile:** |  |  |  |
| Poorest | 3 116/11 412 (27.3) | 1.0 | - |
| Quintile 2 | 3 742/13 365 (28.0) | 1.06 (0.95, 1.18) | 0.33 |
| Quintile 3 | 3 889/15 293 (25.4) | 0.96 (0.86, 1.06) | 0.42 |
| Quintile 4 | 4 087/16 234 (25.2) | 0.98 (0.88, 1.09) | 0.67 |
| Least poor | 4 338/18 047 (24.0) | 0.85 (0.76, 0.95) | <0.01 |
| **Household size:** |  |  |  |
| Multiple residents | 18 851/72 997 (25.8) | 1.0 | - |
| Single resident | 323/1 368 (23.6) | 1.41 (1.14, 1.75) | <0.01 |
| **Household religion:** |  |  |  |
| Islam | 17 976/69 639 (25.8) | 1.0 | - |
| Christian or other | 1 198/4 726 (25.3) | 1.02 (0.85, 1.21) | 0.85 |
| **Minority language:** |  |  |  |
| Chiyao | 18 157/70 620 (25.7) | 1.0 | - |
| Chichewa or other | 1 017/3 745 (27.2) | 1.15 (0.95, 1.39) | 0.16 |
| **Urbanicity:** |  |  |  |
| Rural community | 14 836/55 866 (26.6) | 1.0 | - |
| Peri-urban community | 4 338/18 499 (23.4) | 0.94 (0.74, 1.18) | 0.57 |
| **Treatment (by round):** |  |  |  |
| **cMDA round:** |  |  |  |
| Round 1 | 4 187/11 995 (34.9) | 1.0 | - |
| Round 2 | 4 353/12 285 (35.4) | 0.98 (0.92, 1.05) | 0.58 |
| Round 3 | 2 885/11 930 (24.2) | 0.50 (0.46, 0.53) | <0.001 |
| Round 4 | 2 816/12 296 (22.9) | 0.42 (0.39, 0.45) | <0.001 |
| Round 5 | 2 102/12 538 (16.8) | 0.21 (0.19, 0.23) | <0.001 |
| Round 6 | 2 831/13 321 (21.3) | 0.25 (0.23, 0.28) | <0.001 |
| **Morning household visit(s):** |  |  |  |
| Never early | 18 402/70 985 (25.9) | 1.0 | - |
| Ever early (<9am) | 772/3 380 (22.8) | 1.02 (0.90, 1.15) | 0.75 |
| **Late afternoon visit(s):** |  |  |  |
| Never late | 15 615/59 451 (26.3) | 1.0 | - |
| Ever late (≥3pm) | 3 559/14 914 (23.9) | 0.83 (0.78, 0.87) | <0.001 |

| **Day of visit(s):** |  |  |  |
| --- | --- | --- | --- |
| Never weekend | 18 262/71 002 (25.7) | 1.0 | - |
| Ever weekend | 912/3 363 (27.1) | 0.91 (0.82, 1.00) | 0.08 |
| **Total number of visit(s):** |  |  |  |
| Continuous | - | 0.84 (0.81, 0.88) | <0.001 |

\*: Multilevel mixed-effects logistic regression with 74 351 observations; random intercept at the individual (level-1) and community (level-2) levels, and random slope for CMDA round. †: Defined as an individual who reported to have spent more than half the time away from the household in the six months prior to census.

**Table 3:**
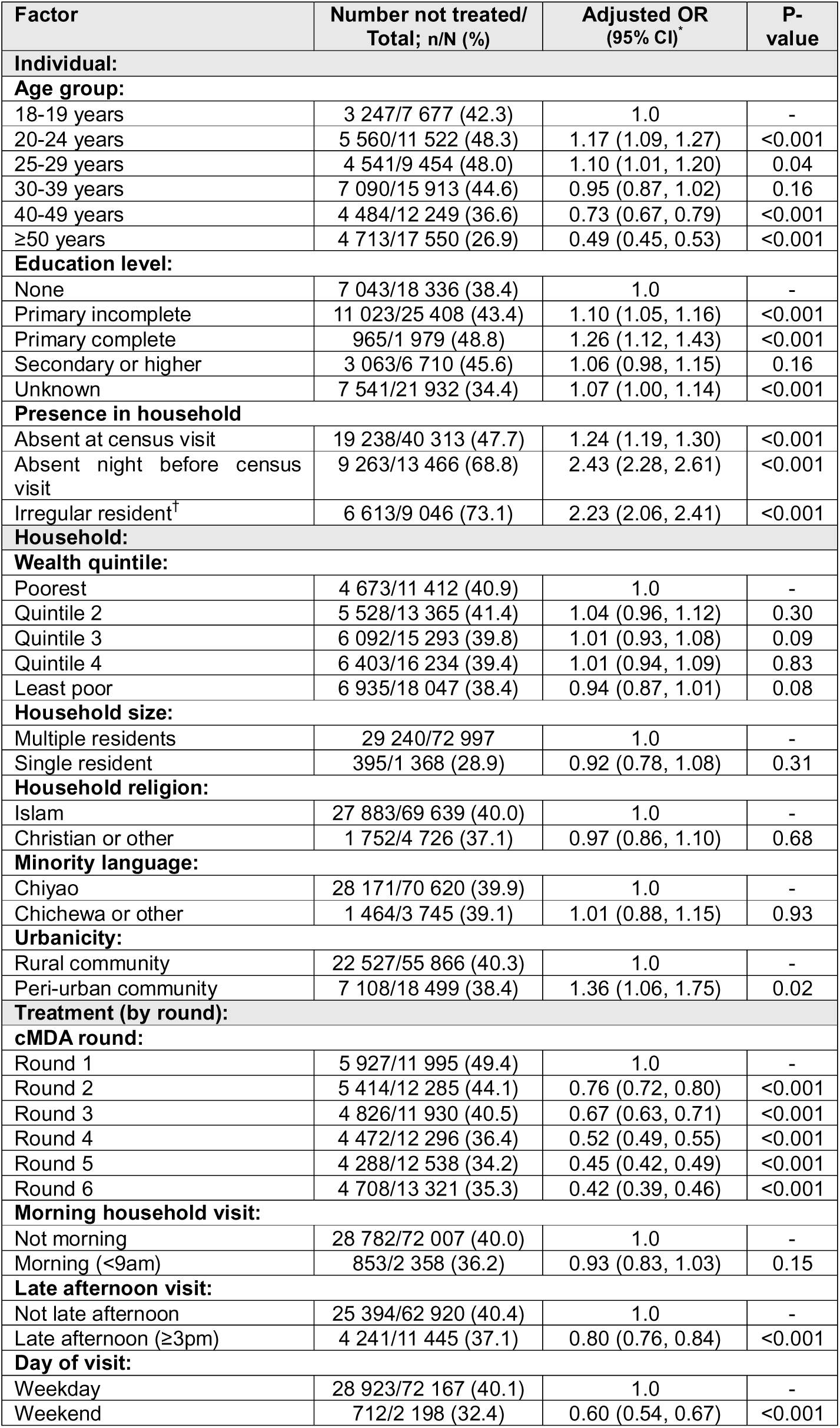
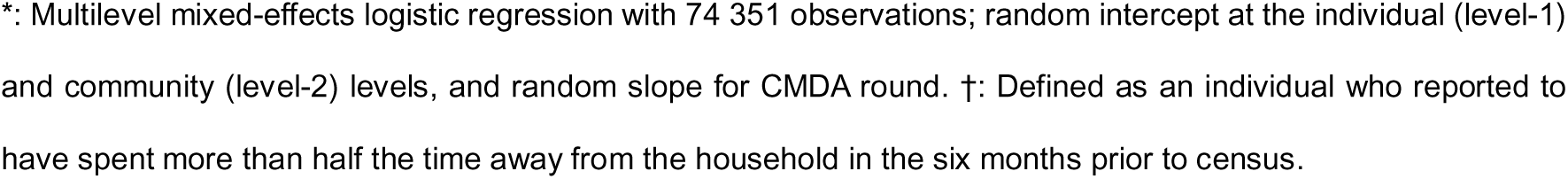
Proportion non-treated and factors associated with operational scenario non-treatment of censused and eligible adult men during cMDA.

Three areas of individual-level characteristics were assessed (age group, education level, and presence in household). Relative to the youngest age group (18-19 years), odds of protocol non-treatment were highest amongst men aged 25-29 years (AOR=1.71, 95% CI 1.52–1.91) and lowest amongst men aged ≥50 years (AOR=0.38, 95% CI 0.34–0.42). Relative to men who had never attended formal education, odds of protocol non-treatment were higher amongst men reported to have completed primary education (AOR=1.19, 95% CI 1.02–1.38); but there was no evidence of an association amongst men with lower (primary incomplete, AOR=1.04, 95% CI 0.97–1.11) or higher (secondary and higher, AOR=1.01, 95% CI 0.91–1.12) education levels. Lastly, increasing levels of reported absenteeism prior to the census enumeration showed a consistent relationship with protocol non-treatment: men who were absent at the time of the census visit (day absent) compared to those who were present (AOR=1.23, 95% CI 1.16–1.30), men reported to have slept elsewhere the night before the census visit (night absent) compared to those who had not (AOR=3.00, 95% CI 2.76–3.25), and men who were reported to have been away from the household for more than half of the time in the past six months (AOR=2.69, 95% CI 2.45–2.96); all had increased odds of protocol non-treatment.

Five areas of men’s household-level characteristics were assessed (wealth, household size, those practicing a minority religion, those primarily speaking a minority language, and those residing in a peri-urban community). Relative to the least wealthy households (quintile 1), odds of protocol non-treatment were lowest amongst men residing in the most wealthy households (quintile 5) (AOR=0.85, 95% CI 0.76–0.95). Men who reported living alone in the previous census visit (single resident household) as compared to those who lived with any other individuals (multiple resident household) showed evidence of an association and greater odds of protocol non-treatment (AOR=1.41, 95% CI 1.14–1.75).

Finally, three areas of treatment-level characteristics were assessed for their association with protocol non-treatment: round of cMDA, times of household visit(s) during cMDA by round, and cumulative number of household visits for cMDA by round. The absolute proportion of men per-protocol not-treated declined round-to-round, from more than one in three in round 1 (34.9% not treated) to one in five in rounds five and six (21.3% not treated). Men’s odds of protocol non-treatment were lowest in round 5 (AOR=0.21, 95% CI 0.19–0.23), and highest in round 1 (reference). In addition, men’s odds of protocol non-treatment overall were consistently lower in cMDA rounds 3-6 as compared to round 2 (AOR=0.98, 95% CI 0.92–1.05). Amongst time characteristics of the visits by each round, men’s odds of protocol non-treatment was significantly lower where any visit to the individual was conducted later in the day (after 3pm) (AOR=0.83, 95% CI 0.78–0.87) (versus never after 3pm). There was no evidence of an association with protocol non-treatment where any visit was conducted earlier in the day (before 9am) (AOR=1.02, 95% CI 0.90–1.15). Finally, there was weak evidence of an association in terms of reduced odds of protocol non-treatment where any visit had been conducted on a weekend (AOR=0.91, 95% CI 0.82–1.00, p=0.08).

### 4.6. Factors associated with non-treatment of adult men: operational scenario coverage

Amongst individual-level characteristics, age group demonstrated a weaker association with operational non-treatment compared to protocol non-treatment; with odds of operational non-treatment also highest amongst men aged 20-24 (AOR=1.17, 95% CI 1.09–1.27) and least amongst men aged ≥50 years (AOR=0.49, 95% CI 0.45–0.53). In contrast, education level demonstrated a stronger relationship with operational non-treatment compared to protocol non-treatment, with men with either primary incomplete (AOR=1.10, 95% CI 1.05–1.16) or primary complete (AOR=1.26, 95% CI 1.12–1.42) both demonstrating increased odds of non-treatment compared to men reporting no formal education; but similarly no evidence was observed of association with operational non-treatment at secondary or higher levels (AOR=1.06, 95% CI 0.98–1.15). Lastly, and consistent with associations observed for protocol non-treatment, increased odds of operational non-treatment were associated with all three measures of household presence, most notably for men absent in the night prior to the census visit (AOR=2.43, 95% CI 2.28–2.61) and men who were irregular residents (AOR=2.23, 95% CI 2.06–2.41).

Amongst household-level characteristics; consistent with protocol non-treatment, there was no evidence of association with operational non-treatment amongst men residing in a household practicing a minority religion (AOR=0.97, 95% CI 0.86–1.10) or speaking a minority language (AOR=1.01, 95% CI 0.88–1.16). There was also no evidence of an association with operational non-treatment amongst men in any household wealth quintile relative to the least wealthy quintile, including within quintile 5 (AOR=0.94, 95% CI 0.87–1.01). However, in contrast to protocol treatment, men residing in a household within a peri-urban community were at increased odds of operational non-treatment (AOR=1.36, 95% CI 1.06–1.75) compared to men residing in a rural community; and conversely, no evidence of an association with operational non-treatment was observed amongst men who lived alone (AOR=0.93, 95% CI 0.79–1.09) compared to men who did not live alone.

Finally, amongst treatment-level characteristics, consistent with protocol non-treatment, men’s odds of operational non-treatment was consistently lower in all rounds of treatment after the first round of cMDA, including round 2 (AOR=0.76, 95% CI 0.72–0.80). Compared to protocol non-treatment, we observed stronger evidence of an association for reduced odds of operational non-treatment when the visit had taken place on a weekend day (AOR=0.59, 95% CI 0.54–0.67) and to a lesser extent in the late afternoon (AOR=0.80, 95% CI 0.76–0.84).

## 5. DISCUSSION

In this study, we show that treatment coverage of adult men in cMDA delivered in the context of a community-based intervention trial was consistently and substantially lower than amongst adult women, with overall protocol treatment coverage of 74.1% of adult men compared to 90.7% of adult women, and a median community-level difference of +15.7% in favour of adult women. When treatment coverage was defined using a pragmatic operational scenario, this difference in coverage substantially increased for adult men, with most communities not achieving ≥75% treatment coverage of adult men despite almost all achieving that benchmark for adult women. While achieving high cMDA coverage amongst women should rightly be recognised as a major achievement overall, this study shows that where cMDA and similar interventions are aiming to achieve high coverage across the general population, it is crucial that sex-related inequities are identified and addressed in both the implementation and evaluation of such strategies. This analysis has explored in detail factors associated with non-treatment of adult men in this setting and highlights challenges in reaching this demographic; including absenteeism, (younger) age, household composition and place of residence, and characteristics of cMDA delivery.

Sex or gender-disaggregated reporting of cMDA coverage remains uncommon, but contemporary evidence supports our finding that adult men’s coverage in cMDA is generally lower than amongst adult women [11, 26]. Amongst studies of routine cMDA programmes that have historically included treatment of adult men (e.g. for onchocerciasis (OC) and lymphatic filariasis (LF)), evidence is fragmented and inconsistent [27], with individual reports of cMDA coverage lower amongst men [28–30] but being equal elsewhere [31, 32] or lower amongst women [33, 34]. A complementary source of evidence for men’s participation in cMDA are studies of non-treatment over time and multiple rounds (i.e. partial or complete non-compliance): amongst four studies that had disaggregated non-compliance by age and sex as identified in a recent systematic review [35], non-compliance was more likely amongst adult men relative to adult women in a study of cMDA for STH in Kenya [36] and for OC and LF in Cameroon [37]; but had no difference in a study of cMDA for trachoma in Ethiopia [38] or OC elsewhere in Cameroon [39]. A key challenge in comparison of cMDA coverage or compliance is the lack of standardised definitions as well as programmatic differences including who is not eligible for cMDA (particularly as it relates to WRA), making interpretation across studies and settings more difficult.

To inform strategies to improve cMDA coverage amongst adult men, we explored a range of demographic and operational factors associated with non-treatment specifically amongst this population. Our results should be interpreted with respect to previous work exploring factors associated with cMDA protocol non-treatment overall amongst all individuals across all three different research sites [22]. We observed greater odds of non-treatment amongst younger men, consistent with the findings of a comparable study of cMDA for STH conducted in Kenya [36]. We also observed that men living alone were at increased odds of protocol non-treatment, highlighting the challenges of reaching such individuals even where resources were optimal. Finally, we observed that men living in peri-urban communities had greater odds of operational non-treatment (but not protocol non-treatment), reflecting the importance of additional efforts required to reach such communities as discussed elsewhere [40].

Short-term absenteeism is inherently a challenge to achieving high coverage of cMDA in general [41–43], and recognition that men are more frequently absent from the household as compared to women – often associated with gender-specific occupational roles and expectations – has previously been described in a variety of settings where cMDA is implemented [12, 44, 45]. In a large cRCT of cMDA for schistosomiasis conducted in Uganda, even where coverage was similar between adult men and women, women were less likely than men to not receive treatment due to absence [46]. This challenge was explicitly identified prior to implementation of cMDA during formative work in the other two DeWorm3 sites: in India, men expressed concerns that drug distribution to households during the day would affect treatment coverage [47] and household-based distribution was identified as a relative benefit to women in Benin in terms of access to treatment [48]. In this analysis, we identified that men who were visited later in the day had lower odds of protocol non-treatment, and men who were visited during the weekend had lower odds of both protocol and operational non-treatment. Household visits ‘out-of-hours’ was not a formal component of cMDA and conflicts with the current scope of role for HSAs [49], but offers a potential lever for programmes in their current form to improve men’s coverage of cMDA.

Understanding men’s coverage in routine cMDA – and sociodemographic differences in coverage of NTD interventions more broadly – remains constrained by a lack of disaggregated reporting as part of routine monitoring and evaluation. Coverage for cMDA has long been assessed through rapid coverage surveys, which generally record a minimum dataset that includes at least sex or gender, age (group) and implementation unit [18]. Despite collection of this data, promotion of reporting and use of disaggregated data is almost entirely absent from current guidelines [50], and sits in contrast with broader calls for disaggregated reporting across the sustainable development goals [51] and WHO adoption of reporting standards for sex and gender [52]. As cMDA strategies are refined in pursuit of more equitable outcomes, and new NTD strategies are developed and tested, it is crucial that greater focus is given to sex and gender disaggregation for monitoring and evaluation, particularly in light of calls for NTD coverage to be an appropriate indicator for assessing progress towards universal health coverage [53].

A key implication of this study is: having recognized and measured differences in cMDA coverage, how can this feasibly be addressed within the current constraints of NTD programmes? Delivery of cMDA in this trial was underpinned by the development of an electronic treatment register to allow real-time monitoring of coverage, a platform that could reasonably be used to target mop-up by sociodemographic characteristics rather than a focus on outright coverage [25]. Research elsewhere can also offer useful guidance to the NTD sector on how to actively engage and incorporate men’s concerns in the delivery of cMDA and other community-health interventions for NTDs, most notably from advances in decentralised HIV testing and linkage to care [54]. A strategy integrating cMDA for schistosomiasis with HIV self-testing for men in Malawi has demonstrated the feasibility of novel approaches to delivering cMDA to men in a more targeted way, and illustrates the ready opportunities for integration of cMDA with other disease-specific and gender-sensitive programmes [55].

This study is subject to several limitations. The implementation of cMDA was conducted in a trial context, with substantial financial and technical investment required to achieve high coverage. While we have attempted to approximate cMDA coverage through an alternative definition that better reflects the realities of routine cMDA delivery, this is likely to be an oversimplification of cMDA delivery and still reflects a substantially better-resourced intervention than many routine programmes. Protocol treatment coverage includes treatments that were not directly observed but instead left with the individual or someone else at the household, and may not have actually been taken, potentially resulting in overestimation of treatment coverage and differential misclassification of the outcome if (for example) men were less likely to comply with treatment when left as compared to women. Finally, we have not explored the effect of cMDA awareness and perceptions of being at-risk of STH, and the role that these factors may play in explaining men’s lower coverage in cMDA, identified as major drivers of coverage in other settings.

## 6. CONCLUSION

The analysis reported here has highlighted the consistently lower coverage of adult men by cMDA implemented in the context of a well-resourced randomised trial, identified factors associated with non-treatment, and more broadly highlighted the importance of exploring socio-demographic differences in the coverage of NTD strategies both operationally and as part of rigorous evaluation.

Amongst the conceptualisations of adult men in global health described by *Beia et al* [56] it is the category of ‘gatekeeper’ that resonates most with research on men’s participation in cMDA to date, with a predominant focus on the role of gender dynamics in access to care: much emphasis has been placed on understanding the role that men may play as intermediaries of women’s participation in cMDA, whether directly as the drug distributor [57] or as husband and head of household [58]. Far less focus has been placed on identifying, exploring and addressing specific challenges men may face in participating in cMDA, a research and operational priority increasingly highlighted as key to achieving genuinely gender-equitable NTD outcomes [59, 60].

We encourage programme managers and implementers looking to achieve better coverage of MDA amongst adult men to consider the implications of the results presented here, but also highlight to policymakers and development partners the critical importance of ensuring that community-level NTD strategies reflect the needs of both men and women.

## 8.1. Acknowledgments

We sincerely thank all community members who participated in DeWorm3 Malawi, and we gratefully acknowledge the support of the traditional authorities of Bwananyambi, Chowe, Jalasi and Katuli; the area development committees of Bwanayambi, Majuni, Malombola and Mandimba, and members of the DeWorm3 Malawi Community Advisory Board.

We thank all those who contributed to DeWorm3 Malawi, with special recognition of the individual contributions of Chikondi Chikotichalera, Fraser Chisale, Hastings Mangawah, Mabvuto Iron, Zachariah Kamwendo, Dickson Mogoya, Cidreck Nkomela and Ranneck Singano.

We thank the team of field enumerators who contributed directly to data collection and the drivers, health surveillance assistants and village volunteers who participated in research activities and the delivery of MDA in Malawi. We also thank those at Mangochi DHO (Dr Henry Chibowa and Dr Kondwani Mamba) and Namwera zone (Mwai Chipeta and Cornelius Kunkeyani) for their support and supervision.

Lastly, we thank Jayne Webster for comments on the first draft of the manuscript, and all those at LSHTM (Katherine Halliday, Eleanor Martins) and University of Washington (Arianna Means, Kristjana Ásbjörnsdóttir, Emily Pearman, Mariyam Shaikh, Katy Sharrock) who contributed to the design, coordination and implementation of DeWorm3.

## 8. ADDENDUM

### 8.2. Financial disclosure

The DeWorm3 study was funded through grants to the Natural History Museum of London (OPP1129535) and University of Washington (INV-022149, INV-030049, INV-002114) from the Gates Foundation. The funder of the study had no role in study design, data collection, data analysis, data interpretation, or writing of this report.

### 8.4. Data availability statement

Access to the data and supporting documents reported in this manuscript is available on request at Vivli (https://doi.org/10.25934/PR00010754).

### 8.5. Author contributions

RLP, RLB, KK & JLW contributed to funding acquisition, conceptualised the parent trial and provided supervision. SWM, JS, RM, AC, ER, JT, HL & LZ contributed to investigation and project administration. SG and WO contributed to software, data curation and validation. SWM conceptualised this study, conducted the formal analysis, generated the figures and wrote the original draft. JS, SRG, WO, ER, HL, JLW, RLP, RLB & KK contributed to the review and editing of the final manuscript.

### 8.6. Patient and public involvement

This study took place under the auspices of the Deworm3 Malawi Community Advisory Board (CAB), whose membership oversees all research activities conducted within the trial site. Members of the CAB were closely engaged in the planning and implementation of the study, but were not directly involved in the design or analysis of the study.

