## Supplementary Tables for "Understanding men’s participation in mass drug administration: evidence from a cluster-randomised trial of community-based deworming in Malawi"

**SUPPLEMENTARY MATERIALS**

**Supplementary table 1: Univariable associations of individual, household and programmatic factors with protocol non-treatment of censused and eligible adult men during cMDA**

| **Factor** | **Unadjusted OR**  **(95% CI)** | **P-value** |
| --- | --- | --- |
| **Individual:** | | |
| **Age group:** | | |
| 18-19 years | 1.0 | - |
| 20-24 years | 2.04 (1.81, 2.28) | - |
| 25-29 years | 2.43 (2.13, 2.77) | - |
| 30-39 years | 1.56 (1.38, 1.77) | - |
| 40-49 years | 0.71 (0.62, 0.81) | - |
| ≥50 years | 0.34 (0.30, 0.38) | <0.001 |
| **Education level:** | | |
| None | 1.0 | - |
| Primary incomplete | 1.11 (1.03, 1.19) | - |
| Primary complete | 1.36 (1.16, 1.61) | - |
| Secondary or higher | 1.30 (1.16, 1.46) | - |
| Unknown | 0.94 (0.85, 1.04) | <0.001 |
| **Presence in household:** | | |
| Absent at census visit | 2.21 (2.08, 2.24) | <0.001 |
| Absent night before census visit | 5.65 (5.30, 6.03) | <0.001 |
| Irregular resident | 6.89 (6.38, 7.44) | <0.001 |
| **Household:** | | |
| **Wealth quintile:** | | |
| Least wealthy | 1.0 | - |
| Quintile 2 | 1.03 (0.89, 1.18) | - |
| Quintile 3 | 0.84 (0.73, 0.96) | - |
| Quintile 4 | 0.81 (0.71, 0.93) | - |
| Most wealthy | 0.70 (0.61, 0.81) |  |
| **Household size:** | | |
| Multiple residents | 1.0 | - |
| Single resident | 0.93 (0.72, 1.21) | 0.61 |
| **Minority religion:** | | |
| Islam | 1.0 | - |
| Christian or other | 1.16 (0.98, 1.38) | 0.09 |
| **Minority language:** | | |
| Chiyao | 1.0 | - |
| Chichewa or other | 1.35 (1.13, 1.62) | 0.001 |
| **Urbanicity:** | | |
| Rural community | 1.0 | - |
| Peri-urban community | 0.89 (0.69, 1.15) | 0.37 |
| **Treatment (by round):** | | |
| **cMDA round:** | | |
| Round 1 | 1.0 | - |
| Round 2 | 0.99 (0.93,1.07) | - |
| Round 3 | 0.51 (0.47, 0.55) | - |
| Round 4 | 0.41 (0.38, 0.45) | - |
| Round 5 | 0.21 (0.19, 0.23) | - |
| Round 6 | 0.26 (0.23, 0.29) | <0.001 |
| **Morning household visit(s):** | | |
| Never morning | 1.0 | - |
| Ever morning (<9am) | 1.04 (0.92, 1.18) | 0.53 |
| **Late afternoon visit(s):** | | |
| Never late | 1.0 | - |
| Ever late (≥3pm) | 0.79 (0.75, 0.84) | <0.001 |
| **Day of visit(s):** | | |
| Never weekend | 1.0 | - |
| Ever weekend | 0.91 (0.81, 1.02) | 0.09 |
| **Number of visit(s) (total):** | | |
| Continuous | 0.88 (0.84, 0.92) | <0.001 |

**Supplementary table 2: Univariable associations of individual, household and programmatic factors with operational non-treatment of censused and eligible adult men during cMDA**

| **Factor** | **Unadjusted OR**  **(95% CI)** | **P-value** |
| --- | --- | --- |
| **Individual:** | | |
| **Age group:** | | |
| 18-19 years | 1.0 | - |
| 20-24 years | 1.38 (1.27, 1.50) | - |
| 25-29 years | 1.38 (1.26, 1.52) | - |
| 30-39 years | 1.12 (1.03, 1.22) | - |
| 40-49 years | 0.77 (0.70, 0.84) | - |
| ≥50 years | 0.43 (040, 0.47) | <0.001 |
| **Education level:** | | |
| None | 1.0 | - |
| Primary incomplete | 1.17 (1.11, 1.24) | - |
| Primary complete | 1.43 (1.26, 1.62) | - |
| Secondary or higher | 1.26 (1.16, 1.37) | - |
| Unknown | 0.97 (0.91, 1.04) | <0.001 |
| **Presence in household:** | | |
| Absent at census visit | 1.90 (1.82, 1.98) | <0.001 |
| Absent night before census visit | 4.31 (4.09, 4.54) | <0.001 |
| Irregular resident | 5.11 (4.80, 5.45) | <0.001 |
| **Household:** | | |
| **Wealth quintile:** | | |
| Least wealthy | 1.0 | - |
| Quintile 2 | 1.04 (0.95, 1.15) | - |
| Quintile 3 | 0.94 (0.87, 1.04) | - |
| Quintile 4 | 0.93 (0.85, 1.02) | - |
| Most wealthy | 0.86 (0.79, 0.94) | <0.001 |
| **Household size:** | | |
| Multiple residents | 1.0 | - |
| Single resident | 0.61 (0.51, 0.73) | <0.001 |
| **Minority religion:** | | |
| Islam | 1.0 | - |
| Christian or other | 0.98 (0.87, 1.09) | 0.68 |
| **Minority language:** | | |
| Chiyao | 1.0 | - |
| Chichewa or other | 1.04 (0.91, 1.18) | 0.60 |
| **Urbanicity:** | | |
| Rural community | 1.0 | - |
| Peri-urban community | 1.34 (1.03, 1.74) | 0.03 |
| **Treatment (by round):** | | |
| **cMDA round:** | | |
| Round 1 | 1.0 | - |
| Round 2 | 0.75 (0.71, 0.79) | - |
| Round 3 | 0.68 (0.64, 0.73) | - |
| Round 4 | 0.53 (0.50, 0.57) | - |
| Round 5 | 0.48 (0.45, 0.51) | - |
| Round 6 | 0.47 (0.44, 0.50) | <0.001 |
| **Morning household visit:** | | |
| Not morning | 1.0 | - |
| Morning (<9am) | 0.95 (0.85, 1.07) | 0.41 |
| **Late afternoon visit:** | | |
| Not late afternoon | 1.0 | - |
| Late afternoon (≥3pm) | 0.81 (0.77, 0.86) | <0.001 |
| **Day of visit:** | | |
| Weekday | 1.0 | - |
| Weekend | 0.63 (0.56, 0.70) | <0.001 |

**Supplementary table 3: Multivariate (adjusted) model parameters (protocol treatment outcome).**

|  | Estimate coefficient (SE) | | | | |
| --- | --- | --- | --- | --- | --- |
| **Parameter** | **Model 1:** | **Model 2:** | **Model 3:** | **Model 4:** | **Model 5:**  **(final)** |
| Fixed-part: |  |  |  |  |  |
| Intercept | -0.738  (0.050) | -0.719  (0.063) | -0.927 (0.070) | 0.407  (0.034) | 0.432  (0.037) |
| Random-part: |  |  |  |  |  |
| Community variance | - | 0.070 (0.016) | - | 0.086 (0.022) | 0.099 (0.025) |
| Individual variance | - | - | 1.948 (0.063) | 1.895  (0.062) | 1.609 (0.070) |
| Slope variance | - | - | - | - | 0.043 (0.005) |
| Deviance | 71603 | 71197 | 68117 | 67917 | 67821 |
| Variance partition coefficients |  |  |  |  |  |
| Community | - | 0.021 (0.005) | - | 0.016 (0.004) | 0.020 (0.005) |
| Individual | - | - | 0.372 (0.008) | 0.376 (0.008) | 0.342 (0.010) |

Model 1: single level model; model 2: two-level variance-components logistic model (level 2=community); model 3: two-level variance-components logistic model (level 2=individual); model 4: three-level variance-components logistic model; model 5: three-level variance-components logistic model with random slope at level 2 (individual). Intercept coefficient is outcome (non-treatment in round), other fixed-effect coefficients not displayed. Deviance (badness of fit) statistic calculated as -2 x log-likelihood. Variance partition coefficient is derived from latent response formulation of the model.
